# An unbiased *de novo* network analysis uncovering causal genes and the developmental intersection between autism and co-occurring traits

**DOI:** 10.1101/2023.04.24.23289060

**Authors:** Catriona J. Miller, Evgeniia Golovina, Joerg S Wicker, Jessie C Jacobsen, Justin M. O’Sullivan

**Author notes:** Corresponding author: Justin M. O’Sullivan.

## Abstract

Autism is a complex neurodevelopmental condition that manifests in various ways. Autism is often accompanied by other neurological disorders, such as ADHD, anxiety, and schizophrenia, which can complicate diagnosis and management. While research has investigated the role of specific genes in autism, their relationship with co-occurring traits is not fully understood.

To address this gap, we conducted a two-sample Mendelian Randomisation analysis and identified four genes located at the 17q21.31 locus that are causally linked to autism in fetal cortical tissue (i.e. *LINC02210, LRRC37A4P, RP11-259G18.1, RP11-798G7.6). LINC02210* was also identified as being causally related to autism in adult cortical tissue. By integrating data from expression quantitative trait loci [eQTLs], genes, and protein interactions we identified that the 17q21.31 locus contributes to the intersection between autism and other neurological traits and conditions in fetal cortical tissue. We also identified an additional distinct cluster of co-occurring traits, including cognition and worry, linked to genetic loci at 3p21.1.

Our results support the hypothesis that an individual’s autism phenotype is partially determined by their genetic risk for co-occurring conditions. Overall, our findings provide insights into the complex relationship between autism and co-occurring conditions, which could be used to develop predictive models for more accurate diagnosis and better clinical management.

## MAIN

Autism is a group of neurodevelopmental conditions exhibiting persistent social communication deficits, accompanied by repetitive sensory-motor behaviours and restricted interests (American Psychiatric Association 2013). Researchers have long associated autism with other conditions. For example, Simonoff et al. (2008) found that 70% of individuals in a stratified subsample of adolescents with autism were found to have at least one co-occurring condition. Common co-occurring conditions reported with autism include ADHD, anxiety, depressive disorders and schizophrenia (Croen et al. 2015; de Lacy and King 2013; Neumeyer et al. 2019; Lai et al. 2019). Thus, it is suggested that autism shares common biological mechanisms with co-occurring phenotypes.

Familial aggregation studies have shown autism to be highly heritable (Gaugler et al. 2014; Pinto et al. 2010; Sandin et al. 2017). Genome-wide association studies (GWAS) and genetic studies have identified that 5-15% of autism cases are explained by chromosomal rearrangements or rare specific single-gene mutations (such as fragile X syndrome) with moderate or high penetrance, while the majority are due to copy number variants (CNVs) and single nucleotide polymorphisms (SNPs) with small individual effect sizes (Pinto et al. 2010; Rodriguez-Gomez et al. 2021; Toma 2020). GWAS studies have identified autism-associated SNPs, including those associated with its co-occurring traits (Sun et al. 2019; Autism Spectrum Disorders Working Group of The Psychiatric Genomics Consortium 2017; Pain et al. 2019).

Previous work by our group has investigated the impact of autism-associated SNPs on the biological pathways underpinning autism (Golovina et al. 2021). However, how they relate to the biological intersection between autism and co-occurring traits, or indeed autism risk itself, is still unclear. The majority of autism-associated SNPs are located within non-coding regions of the genome, consistent with the hypothesis that they mark regulatory regions associated with changes in gene expression (Turner et al. 2016; Yuen et al. 2016). These regions, known as expression quantitative trait loci (eQTLs) spatially interact with their target genes, forming regulatory connections which are either cis- or trans-acting (occurring within < 1 Mb, or > 1 Mb respectively) intrachromosomal, or trans-acting interchromosomal. As regulatory interactions are tissue specific and autism is primarily a neurodevelopmental condition, analysing both fetal and adult brain-specific eQTL information may aid our understanding of the mechanisms through which genetic variants act to increase an individual’s chance of developing autism and its co-occurring traits.

To investigate the interconnectivity of complex polygenic traits, network-based analyses have been previously used by our group (Golovina et al. 2023). In this study, we identified causative and pleiotropic genes that are affected by SNPs associated with both autism and other traits within cortical gene regulatory networks (GRNs) at two different developmental stages. Combining our network analysis with pathway analysis, deep learning of regulatory mechanisms, and finally, comparisons to healthcare data, we identified different clusters of genes and traits (e.g. 17q21.31 linked to neurological traits and 3p21.1 linked to cognition and worry) as being associated with autism and its co-occurring traits.

## RESULTS

### Spatially constrained cortical gene regulatory networks were generated for the fetus and adult

Adult and fetal cortical tissue gene regulatory networks (GRNs) were produced using the CoDeS3D pipeline (Fadason et al. 2018) to analyse common SNPs (minor allele frequency (MAF) ≥0.05) to identify spatially constrained eQTLs within adult (i.e. GTEx eQTL database (GTEx Consortium 2020)) and fetal cortical eQTL datasets (Walker et al. 2019) (Fig. 1). The resulting GRNs were comprised of 580,032 and 1,050,154 spatial eQTLs for the fetal (doi: 10.17608/k6.auckland.22565110) and adult (doi: 10.17608/k6.auckland.22564996) cortical tissue respectively.

**Figure 1:**
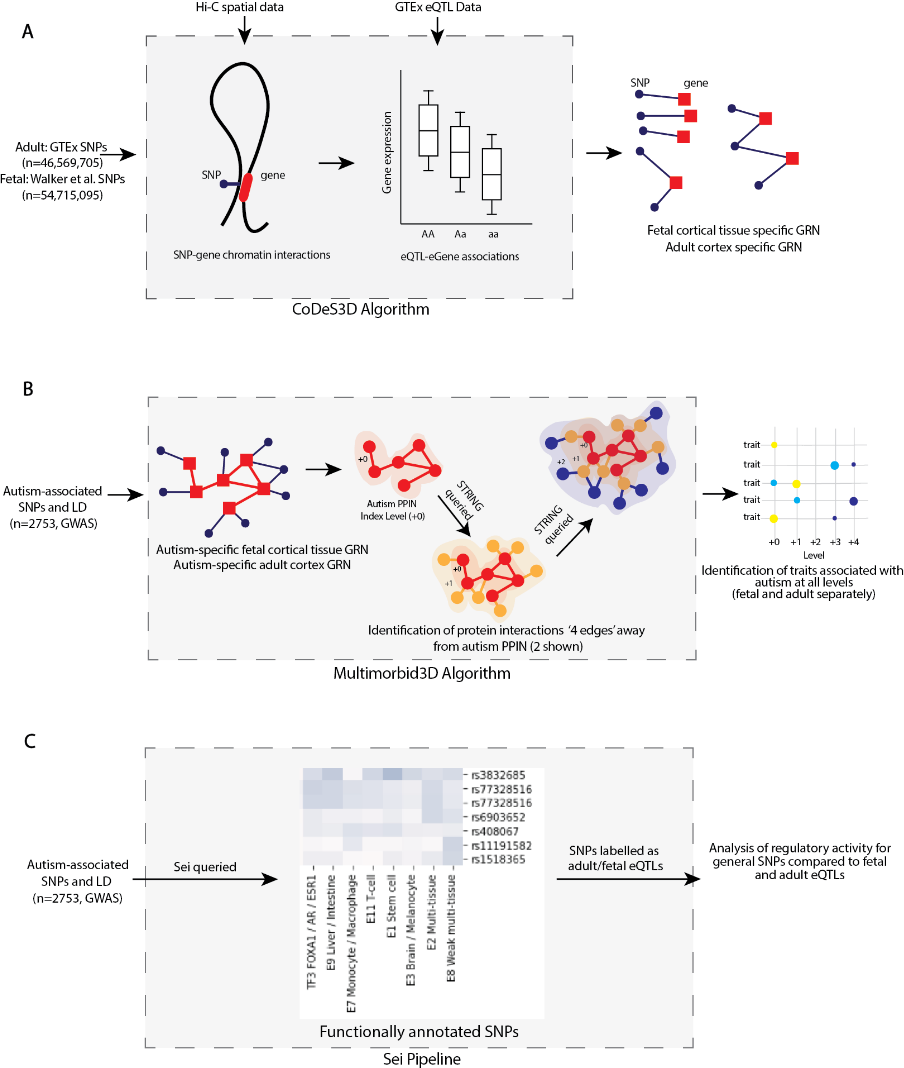
Schematic of methods used in this study. a) Outline of generation of fetal and adult cortical tissue gene regulatory networks (GRN) using CoDeS3D. Hi-C chromatin libraries, derived from fetal brain-specific cortical plate and germinal zone neurons and adult dorsolateral prefrontal cortex cells, were downloaded from dbGaP (accession: phs001190.v1.p1) and GEO (https://www.ncbi.nlm.nih.gov/geo/, accession: GSE87112) respectively. b) The Multimorbid3D algorithm was used to identify traits that co-occur with autism. Autism-associated SNPs (p < 5 x 10^−8^, n = 576; 2753 SNPs including those in LD [r^2^ = 0.8, width = 5000 bp]) from the GWAS Catalog (www.ebi.ac.uk/gwas; 08/05/2022) were used to query the fetal and adult cortical tissue GRNs to generate the autism-specific GRNs. STRING (version 11.5; https://string-db.org) was queried to create a multilevel PPIN for the fetal and adult cortex, separately. c) SNPs were functionally annotated using Sei (Chen et al. 2022). 2753 SNPs, including those in LD with the autism-associated SNPs (supplementary table S2), were queried into Sei. SNPs were collated into LD loci and separated into three groups (loci containing fetal eQTLs, loci containing adult eQTLs, and loci containing only SNPs that were not eQTLs). There was some overlap between the fetal and adult groups.

### Two sample mendelian randomisation identifies four potential causal autism genes within adult and fetal cortical tissue GRNs

A two-sample Mendelian Randomisation (2SMR) study was undertaken using the TwoSampleMR R package (https://github.com/MRCIEU/TwoSampleMR/, version 0.5.6) (Hemani et al. 2018) to identify spatially regulated genes that were potentially causal for autism within the adult and fetal cortical tissue GRNs (supplementary figure 1a). The iPSYCH-PGS 2017 ASD GWAS (Grove et al. 2019) was used as the outcome data. After 2SMR, four genes (*LINC02210, LRRC37A4P, RP11-259G18.1, RP11-798G7.6*) were identified as being statistically significant (Bonferroni adjusted p-value < 0.05) and having a causal role in autism within fetal cortical tissue (supplementary figure 1b). Within the adult cortical tissue GRN, only *LINC02210* was identified as being statistically significant.

### Fetal and adult protein-protein interaction networks identify traits that are pleiotropic with autism

The fetal and adult cortical GRNs (Fig. 1a) were queried with autism-associated SNPs (GWAS Catalog [https://www.ebi.ac.uk/gwas/], p ≤ 5 × 10^−8^; supplementary table S1) and those within LD (r^2^ = 0.8, width = 5000 bp; supplementary table S2) to identify eQTL-gene associations and create autism-specific GRNs (Fig. 1b). The adult autism-specific GRN consists of 888 cis-acting, 63 trans-acting intrachromosomal, and 5 trans-acting interchromosomal eQTL-gene pairings. The fetal network contains 1155 cis-acting, 26 trans-acting intrachromosomal and 4 trans-acting interchromosomal eQTL-gene regulatory connections.

The STRING database was used to create protein-protein interaction networks (PPINs) that extended the autism specific GRNs by four levels, where the proteins on each level interact with a protein encoded by a gene on the previous level (Fig. 1b).

Querying the GWAS Catalog with the eQTLs that associated with genes encoding proteins from each level of the network identified traits that are associated with autism (Fig. 2a; supplementary figures 2 and 3; supplementary table S3). Due to the developmental nature of autism (American Psychiatric Association 2013), we hypothesised that there would be a mixture of shared and unique traits identified in the fetal and adult PPINs. Across the five levels (level 0 – level 4), 44 traits were shared (52% of the adult network and 56% of the fetal network) (Fig. 2b). A higher proportion of the index level traits (level 0) were shared between the fetal and adult PPIN than the outer level traits (p < 0.0001). There were 63 significant (hypergeometric test p < 0.05) traits on level 0 of the adult PPIN and 57 on level 0 of the fetal PPIN, 39 of which appeared in both (Fig. 2c). The finding that the majority of the shared traits were on the index level of the PPIN is notable. The eQTLs for these level 0 traits target genes whose transcript levels also correlate with autism associated eQTLs. Thus, these level 0 genes are pleiotropic with autism.

**Figure 2:**
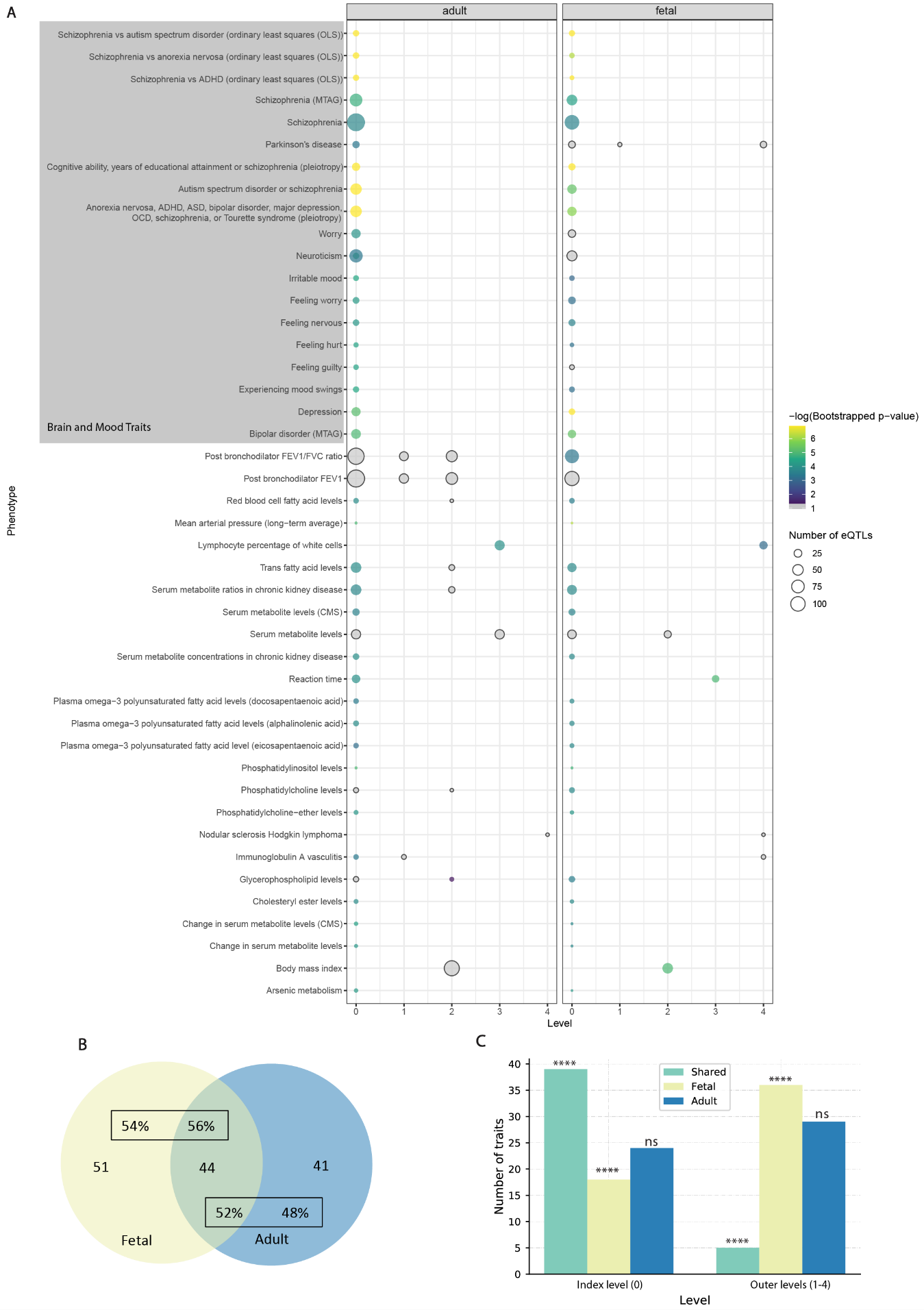
Comparing traits identified during the multimorbid3D analysis for the fetal and adult group. a) Traits identified during the multimorbid3D analysis shared between the adult and fetal group using the STRING database to expand the PPIN. Some traits appear on multiple levels. Circle size indicates the number of eQTLs whilst colour indicates the negative logged p-value from bootstrapping. Y-axis labels are based on GWAS trait names. Traits which passed the hypergeometric test (p < 0.05) but were insignificant (p ≥ 0.05) after bootstrapping are shaded grey. Note: neuroticism has also been misspelt as neurociticism in the GWAS Catalog meaning it appears twice. b) Venn Diagram outlining shared and unique traits across the autism PPIN (level 0-4). 44 traits appeared in both groups, representing 56% of the fetal traits and 52% of the adult traits. c) Bar graph outlining the number of traits identified at different levels of the PPIN. Graph shows that the majority of shared traits are at level 0 (**** = p-value < 0.0001 for shared traits and fetal traits. No difference in adult traits between index and outer levels; ns = not significant p-value = 0.179).

**Figure 3:**
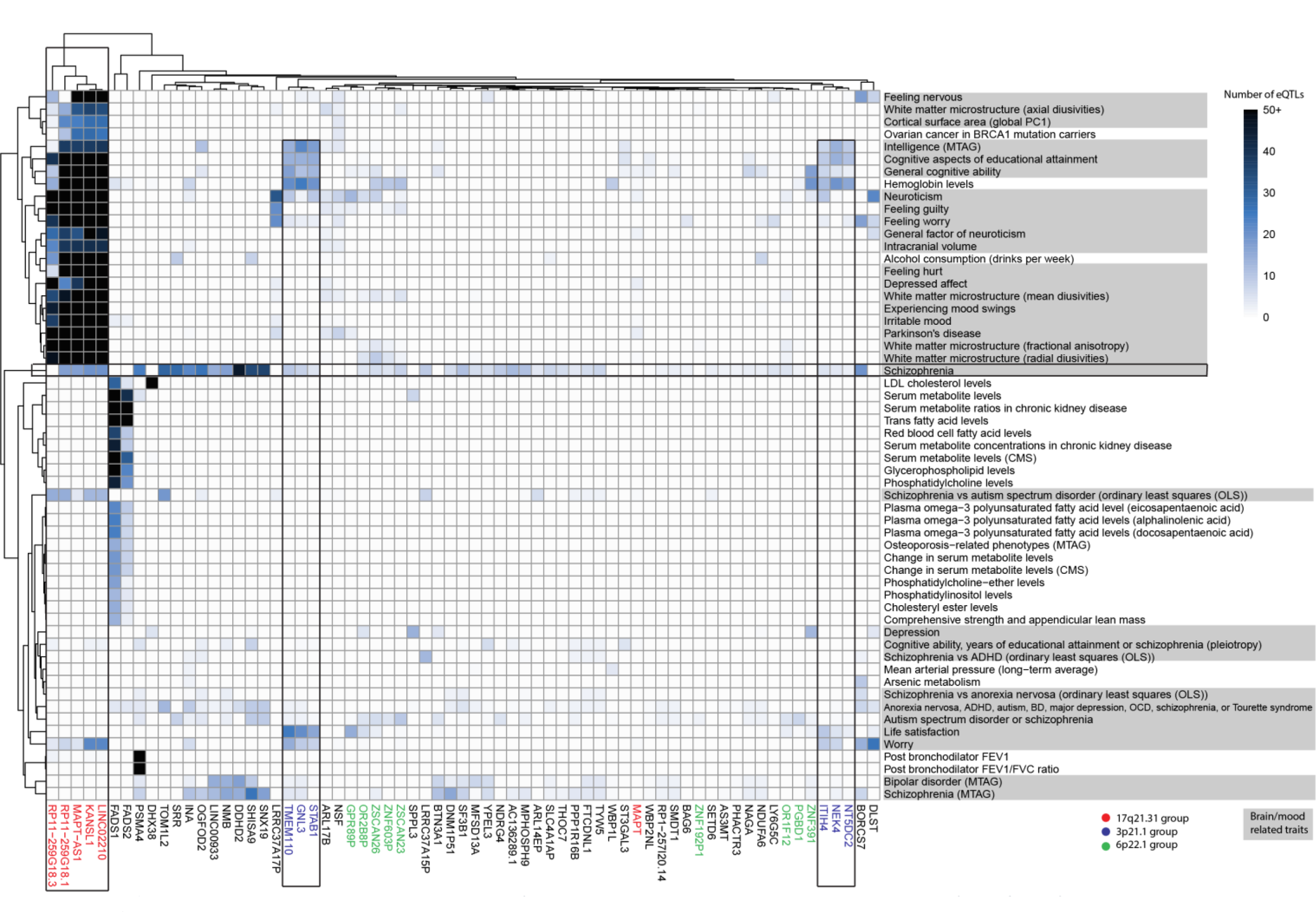
De novo network analysis identified gene clusters and pleiotropic traits in the fetal GRN for autism. Bi-clustering was performed on the gene trait associations using eQTL frequency to identify clusters. eQTL–gene data was obtained using multimorbid3D. Brain and mood related traits are shaded grey. Text coloured based on gene clusters. Black outlines indicate – the chromosome 17 genes (KANSL1 – RP11-259G18.1) relating to a range of mood and brain traits, the chromosome 3 groups (TMEM110 – STAB1 and ITIH4 – NT5DC2) relating to cognition and worry, and the set of genes at the intersection of autism and schizophrenia. See supplementary table S4 for raw output.

Of the index level traits identified in the fetal PPIN, 54% were brain or mood related (Fig. 3). Many of the eQTLs that were responsible for the associations correlated with the transcript levels of genes located at chromosome 17q21.31, including *LINC02210* and *RP11-259G18.1* which were identified as being causal for autism. Notably, of the genes located in 17q21.31, only *KANSL1* was represented on the index level of the adult PPIN (Fig. 4). Other clusters of genes encoding proteins present within the fetal PPIN included (supplementary table S4): 1) metabolic traits associated with *FADS1/2*; 2) traits linked with cognition and worry that associated with two gene clusters (*TMEM110*, *GNL3*, *STAB1* and *ITIH4*, *NEK4*, *NT5DC2*) located at chromosome 3p21.31; and 3) multiple genes across different locations associated with schizophrenia (Fig. 3). Whilst proteins encoded by other genes are present, these groups provide a clear separation of the main traits that appear on the index level of the fetal PPIN.

**Figure 4:**
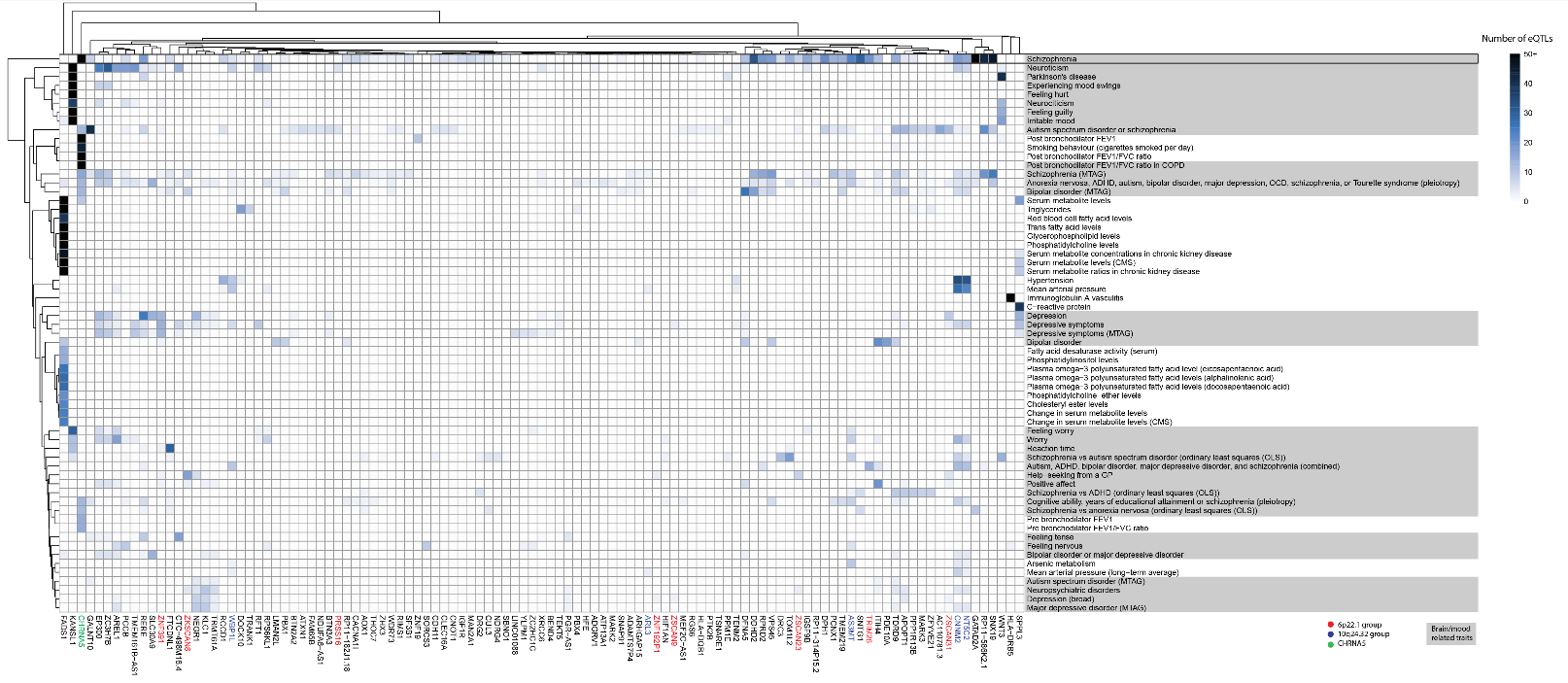
De novo network analysis identified autism associated gene clusters and pleiotropic traits within the adult cortex. Bi-clustering was performed on gene trait associations using eQTL numbers. Data was derived from a multimorbid3D analysis of the adult cortical GRN. For simplicity, all rows with only one eQTL-gene connection were removed (see supplementary table S5 for raw output).

The index level of the adult PPIN contained 52% brain or mood related traits (Fig. 4). Of the non-neurological traits, there were multiple lung related traits linked to eQTLs associated with *CHRNA5* (supplementary table S5). Contrary to the fetal PPIN, there was no detectable clustering of gene loci. However, loci at chromosome 6p22.1 and 10q24.32 were still associated with autism and co-occurring conditions in the adult cortex. Similar to observations of the fetal PPIN, multiple genes from different chromosomal locations were identified as falling at the intersection between schizophrenia and autism.

### Neurological and non-neurological traits were enriched on outer levels of the adult and fetal protein interaction networks

We identified neurological and non-neurological traits as being enriched for the eQTLs associated with the transcript levels of the genes encoding the interacting proteins present in the outer levels of the PPIN (supplementary figures 4 & 5; supplementary table S4 & S5). BMI associated eQTLs were correlated with transcript levels from genes encoding proteins in both the fetal and adult PPINs on level 2 (17 and 57 genes respectively). Notably, eQTLs on the index and outer levels, of both the fetal and adult PPINs, were associated with Parkinson’s Disease. Across both PPINs, eQTLs affecting multiple genes at different levels were associated with traits related to plasma fatty acid and serum metabolite levels. Immune-related traits (e.g. white blood cell count) were also associated with eQTLs for genes encoding proteins on the outer levels of both the adult and fetal PPINs.

Pathway enrichment analysis identified 4 and 26 biological pathways in the fetal and adult PPINs, respectively, that were shared between the index and outer protein interaction levels (i.e. level 0 and levels 1-4) (supplementary table S6). Many of these pathways (2 fetal and 14 adult) were disease-related (e.g. asthma, rheumatoid arthritis and type 1 diabetes mellitus) while other pathways were immune or signalling related (supplementary table S6).

### Deep learning provides insight into regulatory elements associated with autism

The autism-associated SNPs and those in LD (n=2753) were queried for patterns of known regulatory elements using Sei (Chen et al. 2022) – a deep learning algorithm for predicting regulatory activity (Fig. 1c). The Sei deep learning sequence model scored each SNP across 40 different regulatory element patterns, 12 of which referred to enhancer activity (supplementary table S7). SNPs were clustered using LD (r^2^ = 0.8, width = 5000 bp; supplementary table S2) into 60 fetal and 113 adult eQTL containing loci, and 290 loci containing non-eQTL SNPs (i.e. GWAS SNPs that were not present in the fetal or adult GRNs; supplementary figure 6a). Across these autism-associated genetic loci, Sei predicted that the fetal and adult eQTL containing loci were enriched for enhancers, compared to the 290 loci containing only non-eQTL SNPs (supplementary figure 6b). When looking at the highest scoring regulatory class for each SNP, the fetal and adult eQTLs had a significantly higher mean score than the non-eQTL SNPs (supplementary figure 6c). Notably, there were no significant differences between the adult and fetal eQTL containing loci for enhancer enrichment or mean Sei scores (supplementary figure 6b & 6c).

The fifty SNPs with the highest Sei regulatory scores (supplementary figure 7) typically scored highest in either: 1) the erythroblast-like, multi-tissue, or weak epithelial enhancer categories; 2) the promoter category; or 3) the CTCF-Cohesin category. The top 50 SNPs were evenly split between those that were predicted to increase or decrease regulatory activity when compared to the reference sequence at that position. This finding was also true for the top 50 SNPs in the adult and fetal groups (i.e. loci containing at least one eQTL from the adult and fetal GRNs respectively; supplementary figures 8 and 9). The adult group eQTL (rs308107) that had the highest predicted regulatory score (i.e. 7.93) was associated with *RERE* transcript levels. rs308107 was associated with increases in regulatory activity in enhancer, promotor, and CTCF-Cohesin categories (supplementary table S7). rs308107 has been associated with the intersection between major depression (MD) and intelligence, and expression of *RERE* in the brain (Bahrami et al. 2021). *RERE* is a syndromic autism gene (SFARI score S1; https://gene.sfari.org/database/human-gene/RERE) and mutations here have been associated with autism (Fregeau et al. 2016).

### Traits predicted to co-occur with autism are present in Autistic New Zealanders

ICD-10 codes from hospital discharge data were used to calculate odds ratios (ORs) for all conditions that co-occur with autism within the population of autistic New Zealanders who had been admitted to hospital between 2015-2020 (n=2,622; supplementary table S8). Those conditions that were significant (Fisher exact test; p ≤ 0.05) when compared to the general hospitalised population in the same time-period (n=2,051,658), following multiple testing correction, were retained and those that overlapped the traits detected by Multimorbid3D were identified (Fig. 5). Neurological conditions such as schizophrenia, mood disorders, and depression showed an increased prevalence in autistic New Zealanders (i.e. log_10_(OR) > 0). These traits were associated with chromosome 17q21.31 in our Multimorbid3D analysis. However, Parkinson’s disease was not significantly associated with autism within the New Zealand population. Notably, Alzheimer’s disease showed a decreased prevalence in the autistic population. These findings may be due to a smaller number of autistic individuals at an age where Parkinson’s disease and Alzheimer’s disease generally occurs due to recent increases in diagnostic testing (Fombonne 2009). Alzheimer’s disease appeared as a disease pathway in our pathway analysis within the fetal gene group. Whilst it wasn’t present in the adult group, ‘pathways of neurodegeneration’ was present (supplementary table S6). Abnormal weight gain was associated with over 40 genes on the outer levels of the adult PPIN and had an increased OR in autistic New Zealanders. COPD occurred at reduced rates in autistic New Zealanders, and while it was not directly identified in our Multimorbid3D analysis, many lung-related phenotypes including ‘post bronchodilator FEV1/FVC ratio in COPD’ appeared on both the index and outer layers of the adult PPIN (Fig. 4).

**Figure 5:**
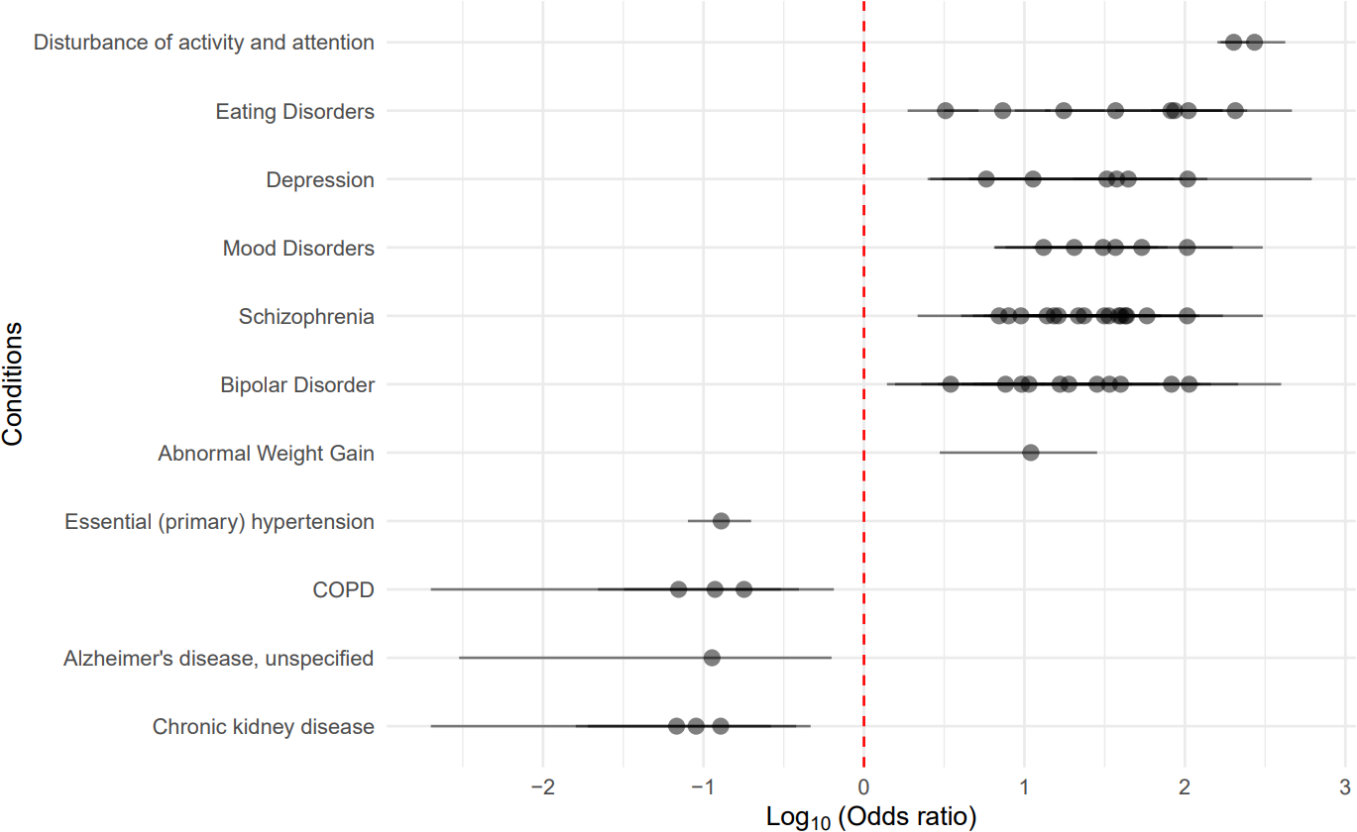
Odds ratios for conditions identified as co-occurring within autistic New Zealanders. Conditions that occurred at significantly (p < 0.05 following multiple testing correction) different rates, when compared to the hospitalised New Zealand population, and overlapped traits identified by Multimorbid3D (i.e. *figure 3* and 4) are shown. Many conditions had multiple ICD-10 codes (e.g. childhood autism and Asperger’s Syndrome for autism and multiple subcodes for schizophrenia).

## DISCUSSION

We performed a *de novo* network analysis of cortical gene regulatory networks that identified four genes that are causal for autism within the 17q21.31 loci in fetal tissue (*LINC02210, LRRC37A4P, RP11-259G18.1, RP11-798G7.6),* of which *LINC02210* was also identified as being causally related to autism in adult tissue. Our analysis identified genetic variants that are associated with traits that are known to co-occur with autism (e.g. schizophrenia (Krieger et al. 2021; Hsu et al. 2022; Chien et al. 2021) and BMI (Croen et al. 2015)). We contend that the results of this study provide a starting point for the individualized stratification of the biological mechanisms linking autism and its co-occurring traits.

The nature of the datasets that were used in this work result in several limitations (Tam et al. 2019). 1) There is an inherent bias towards common traits that are more ‘popular’ to study by GWAS; and 2) not all genetic variants will be represented as GWAS participants to date have been primarily of European ancestry. Moreover, the implemented methods assume the proteins encoded by the target genes form characterised protein-protein interactions. Therefore, any regulatory connections that associate with genes encoding proteins that do not form, or form unknown, protein-protein interactions will not be identified. Furthermore, transcript-levels alone are insufficient to explain protein expression (Battle et al. 2015; GTEx Consortium 2020; Brion et al. 2020). For example, post-transcriptional processes contribute to expected protein levels (Brion et al. 2020; Battle et al. 2015). Therefore, we must be careful when assuming that there is a direct relationship between changes in eQTL associated transcript levels, protein expression and phenotypes. Despite these limitations, the spatially constrained gene regulatory networks we have described can be used to identify 1) causal genes, 2) the traits and conditions that co-occur with autism, 3) the genetic risk that links them, and 4) the potential mechanisms responsible for the observed associations. Collectively, these results provide a step change towards understanding how genetic variation contributes to autism and brings us closer to stratification for diagnosis and long-term management according to an individual’s combination of co-occurring traits.

The individualistic nature of autism is supported by our findings that link variation in distinct genetic loci (SNPs, genes and chromosomal regions) to ‘clusters’ of traits. A similar observation was made by Fu et al. (2022), who found that a distinct subset of genes contributes to the genetic overlap between autism and developmental disorders, when compared to autism and schizophrenia. Our study also supports this notion, as we found that the overlap between autism and other traits may involve subsets of genes that are specific to the overlap of interest.

Within the fetal PPIN, a common group of genes (i.e. *KANSL1*, *LINC02210*, *RP11-259G18.3*, *MAPT-AS1*, *RP11-259G18.1*, and *MAPT*) were associated with the majority of the mood/brain related phenotypes (e.g. neuroticism, Parkinson’s disease, depression, mood swings, and white matter microstructure). Many of these phenotypes, particularly depression and mood traits (Chien et al. 2021; Virues-Ortega et al. 2017; Croen et al. 2015), are known to co-occur with autism. Consistent with this, depression and mood disorders were observed to have an increased prevalence in autistic New Zealanders. The genes that associate with these trait intersections are located within chromosome 17q21.31. The 17q21.31 locus is inverted in ∼20% of Europeans and has previously been reported as an autism susceptibility locus (Pain et al. 2019). There is also a microduplication syndrome at this locus linked to autistic features (Grisart et al. 2009). Notably, we have identified that the regulatory impacts on the genes within this locus involve genes that are causally related to autism and occur in fetal cortical tissue – not the adult cortex. The known functions of the proteins encoded by genes within 17q21.31 provides possible insights into this relationship, albeit not the causal genes which are non-coding or pseudogenes. For example, *MAPT* regulates the expression of Tau, potentially resulting in the development of tauopathy in the brain (Grigg et al. 2020). An increase in tauopathy has been identified in post-mortem brain sections from an autistic child (Grigg et al. 2020). High *MAPT* mRNA levels have also been identified in fetal stages of the frontal cortex when compared to similar samples from childhood and adulthood (Xie et al. 2021). *KANSL1* has already been identified as having a syndromic causative role in autism with a SFARI score of S1 indicating the gene has been clearly implicated in autism (https://gene.sfari.org/database/human-gene/KANSL1). Koolen-de Vries syndrome, partially characterised by developmental delays and behavioural features, is caused by microdeletions or loss of function mutations in *KANSL1* (Koolen et al. 2016). With respect to the causally related genes, *RP11−259G18.1* has been observed to be upregulated in the fetal brain (Pain et al. 2019). However, neither *LINC02210* nor *RP11−259G18.1* have previously been identified as causal autism genes.

A gene cluster (i.e. *NEK4*, *GNL3*, *NT5DC2*, *TMEM110*, *STAB1*, and *ITIH4*) located at chromosome 3p21.1 was identified as associating with co-occurring traits within the fetal PPIN. Proteins encoded by three genes (i.e. *RFT1*, *ITIH4*, and *TMEM110*) within 3p21.1 were also present within the adult PPIN. Multimorbid3D identified that the genetic variants affecting gene expression within the chromosome 3p21.1 locus are involved in the intersection between autism and cognition, worry, and intelligence. Notably, Summary-based Mendelian Randomisation and numerous GWAS studies (Yang et al. 2020; Eum et al. 2021; Burmeister et al. 2009) have associated chromosome 3p21.1 with psychiatric disorders^21–23^. However, we did not observe a causal relationship within the fetal and adult cortical tissues. Links with autism are largely restricted to case and family studies of *NT5DC2*, which provide conflicting results on the gene’s significance (Xie et al. 2020; Wang et al. 2019). Overexpression of *NEK4* and *GNL3* in mice has been linked to reduced mushroom spine density in neurons, which has been associated with cognition and memory (Yang et al. 2020).

Whilst there was a difference in gene clusters and traits affected by spatially constrained eQTLs within the fetal and adult cortical tissue, there were also notable commonalities. For example, Multimorbid3D identified a large genetic intersection between autism and schizophrenia. The connection between autism and schizophrenia has been observed in epidemiological studies (Krieger et al. 2021; Hsu et al. 2022; Chien et al. 2021). Consistent with this, we identified an increased prevalence of schizophrenia in autistic New Zealanders. Previous studies have suggested there is a range of genetic variants at the intersection of these traits (Moreau et al. 2021; Chen et al. 2021; Rees et al. 2021). We identified 54 and 73 pleiotropic genes linking these phenotypes within the fetal and adult cortical tissues, respectively. These pleiotropic genes were distributed throughout the genome and the trait intersections were typically associated with a restricted number of eQTLs (N_fetal_ = 2.6 ± 1.6, N_adult_ = 2.3 ± 1.6). Notably, of the 50 eQTLs predicted by Sei to have the highest regulatory scores in the fetal and adult groups, 42% and 58% were associated with schizophrenia respectively. In particular, *BORCS7*, *INA*, and *SRR* were associated with eQTLs which scored highly for reduced enhancer, promotor and CTCF-cohesin activity. Polygenic risk score (PRS) for schizophrenia has been shown to be predictive in autism (Antaki et al. 2022). However, here we have identified the intersection, not simply the PRS but both the genetic risk and the possible mechanistic link. It also demonstrates that schizophrenia is linked to a subset of the possible genetic variation associated with autism, not every gene. Collectively, these findings are consistent with a complex biological link that raises the chances of some autistic individuals exhibiting features of schizophrenia.

There was reduced clustering of traits – and genes – within the PPIN produced from the adult cortex, possibly reflecting the fact that autism is childhood onset. This relates to our finding of only one causal autism gene (*LINC02210*) within the adult cortical tissue, compared to the four causal genes within the fetal tissue. Despite this, we identified a strong association with respiratory related phenotypes, particularly on level 0 (i.e. *CHRNA5*) and 1 (i.e. *CHRNA3*, *PSMA4*, *HYKK*). Of these, *PSMA4* was a pleiotropic gene within the fetal PPIN. *CHRNA5*, *CHRNA3*, *PSMA4*, and *HYKK* are located at 15q25.1, a locus that is associated with chronic obstructive pulmonary disease (COPD) (Nedeljkovic et al. 2018). Over 50 eQTLs were associated with *CHRNA5* which was associated with post bronchodilator FEV1 and FEV1/FVC ratio. KEGG Pathway analysis also identified asthma as a pathway associated with the index and outer levels of the adult PPIN. Links between autism and lung-related phenotypes have rarely been documented but one case-study demonstrated that a group of 49 autistic children had abnormal lung anatomies when compared to 410 neurotypical control subjects (Stewart and Klar 2013). However, no further validation of this study (Stewart and Klar 2013) has occurred and studies on connections between autism and asthma show mixed results (Zerbo et al. 2015; Kotey et al. 2014; Gong et al. 2022). Despite this, SNPs associated with *CHRNA5* have been associated with a decrease in connectivity of a cortical output circuit linked with a cognitive profile that includes deficits in attention seen in autism (Bailey et al. 2012). We observed a decreased prevalence of COPD within autistic New Zealanders, suggesting autism is protective against COPD. Thus, whilst our study identifies a potential connection between autism and COPD, it is important to remember that gene regulation is tissue specific, and our analysis was run on cortical tissue GRNs.

Parkinson’s and Alzheimer’s disease were linked to autism through protein interactions (and thus eQTL – gene associations) that appeared on the outer level of the fetal PPIN (Alzheimer’s disease) and index and outer levels (Parkinson’s disease) of both the fetal and adult PPINs. These neurodegenerative conditions were also identified as being enriched in the fetal pathway analysis, while ‘pathways of neurodegeneration - multiple diseases’ was enriched in the adult analysis. Parkinson’s disease was associated with the 17q21.31 loci within the fetal analysis, including *LINC02210* and *RP11-259G18.1* which were identified as causal autism genes. Parkinson’s disease has been identified as being significantly more common in autistic individuals^4^. Alzheimer’s disease, Parkinson’s disease, and autism have all been associated with neuronal nicotinic acetylcholine receptors (Henderson et al. 2012). Within autistic New Zealanders, we saw a reduced prevalence of Alzheimer’s disease and no statistically significant relationship between autism and Parkinson’s disease. However, the analysis of autistic New Zealanders did not adjust for age and thus the apparent contradiction may be due to a smaller number of autistic individuals at an age where Parkinson’s disease and Alzheimer’s disease generally occurs. Most standardised diagnostic testing for autism wasn’t available until the 21^st^ century, meaning many adults who would now display ageing diseases would have been unlikely to receive a diagnosis (Fombonne 2009).

The findings from our network and epidemiological analyses provide insights into the time-dependency of the genetic mechanisms involved in the interaction between autism and co-occurring traits. We have highlighted ‘developmental windows’ (e.g. fetal development) where genetic loci (e.g. 17q21.31 and 3p21.1) have the potential to impact on these trait combinations. This is supported by the identification of causal autism genes within the 17q21.31 region. It has previously been suggested that autism does not operate on a linear schedule, but is influenced by a combination of factors (Antaki et al. 2022). In this study we provide a biological argument for looking at an individual’s phenotype as being related to their combined genetic risk for different clusters of traits. Future work should test the utility of these genetic associations to create predictive models to stratify individuals for diagnosis and long-term management of the co-occurring conditions that confound autism and impact on each individual’s quality of life.

## MATERIALS AND METHODS

### Creation of the fetal and adult cortical tissue gene regulatory networks

The CoDeS3D algorithm (Fadason et al. 2018) (https://github.com/Genome3d/codes3d) was used to develop gene regulatory networks (GRN) for fetal and adult cortical tissue. All SNPs present in the adult cortical tissue specific GTEx (GTEx Consortium 2020) eQTL database (n = 46,569,705) and those from the fetal dataset (Walker et al. 2019) (n = 54,715,05) were input into CoDeS3D.

Hi-C chromatin libraries, derived from fetal brain-specific cortical plate and germinal zone neurons (Won et al. 2016) and adult dorsolateral prefrontal cortex cells (Schmitt et al. 2016), were downloaded from dbGaP (accession: phs001190.v1.p1) and GEO (https://www.ncbi.nlm.nih.gov/geo/, accession: GSE87112) respectively.

This was used to generate a list of spatial SNP-gene pairs based on interactions between restriction fragments containing the queried SNPs and restriction fragments overlapping genes. From this, the adult (GTEx Consortium 2020) and fetal (Walker et al. 2019) cortical tissue eQTL datasets were used to determine which SNPs were eQTLs (Fig. 1a). Benjamini Hochberg multiple testing was performed to identify significant (adjusted p-value ≤ 0.05) pairings.

### Two Sample Mendelian Randomisation

To identify any genes potentially causal for autism within the adult and fetal cortical tissue GRNs, a two sample mendelian randomisation (2SMR) study was completed for adult and fetal separately (supplementary figure 1a). This was done using the TwoSampleMR R package (https://github.com/MRCIEU/TwoSampleMR/, version 0.5.6) (Hemani et al. 2018). The eQTL-gene pairs within each adult and fetal cortical tissue GRN (output of figure 1a) were used as the exposure instruments. All eQTLs with an exposure p-value > 1 × 10^−5^ were also removed from the exposure dataset. Clumping was then undertaken to ensure all exposure instruments were independent (Hemani et al. 2018). The iPSYCH-PGS 2017 ASD GWAS (Grove et al. 2019) was chosen for the outcome data due to its size. This was downloaded within the TwoSampleMR package from the IEU Open GWAS Project (Elsworth et al.). After harmonising the exposure and outcome data, genes with one eQTL associated with it underwent 2SMR using the Wald test whilst those with multiple eQTLs underwent 2SMR using MR Egger regression and inverse variance weighted methods (Dang et al. 2022). A Bonferroni correction was used to adjust the p-value threshold to correct for multiple tests. Genes whose p-values were above this threshold were considered statistically significant 2SMR results and therefore have a causal role in autism within adult or fetal cortical tissue.

### Definition of autism-associated SNPs

A keyword search for the exact term “Autism” identified 576 autism-associated SNPs (*p* < 5 × 10^−8^) in the GWAS Catalog (supplementary table S1). These SNPs were downloaded (www.ebi.ac.uk/gwas; 08/05/2022).

### Identifying possible co-occurring traits of autism

Potential co-occurring traits were identified using the Multimorbid3D pipeline (Golovina et al. 2023; Zaied et al. 2023) (https://github.com/Genome3d/multimorbid3D). The 576 autism-associated SNPs underwent a linkage disequilibrium analysis to determine those in LD (r^2^ = 0.8, width = 5000 bp; supplementary table S2). 2753 SNPs (n = 576 + LD) were input into Multimorbid3D which queried the fetal or adult cortical tissue specific GRN created in figure 1a to create an autism specific fetal or adult GRN. Proteins encoded by the genes in this GRN formed the ‘Level 0’ network. Protein interactions proximal to the autism-associated network (under 4 ‘edges’ away; where an edge refers to a direct interaction) were identified using STRING (https://string-db.org; Fig. 1b). Interactions came from experiments, text mining, co-expression and databases, and was limited to species “Homo sapiens” and an interaction score above 0.7 (Szklarczyk et al. 2019).

The ‘Level 1’ proteins were those that were identified as interacting directly with the autism-associated proteins, whilst level 2-4 proteins were those interacting with proteins on the previous level. At each level, the genes encoding the proteins were used to query the fetal or adult cortical tissue GRNs to identify their associated significant regulatory eQTLs (adjusted p-value ≤ 0.05). These eQTLs at each level were queried against the GWAS Catalog to identify traits associated with them (hypergeometric test; p<0.05). This process was completed for both fetal and adult cortical tissue separately to create the fetal and adult protein-protein interaction networks (PPINs).

### Bootstrapping analysis

Following the Multimorbid3D analysis, 1000 bootstrapping iterations were performed to mitigate any errors from the traditional sampling approach assumption that the sampling distribution will be approximately normal. At each iteration, 2753 trait-associated SNPs were randomly selected from the GWAS Catalog and run through the Multimorbid3D pipeline. After 1000 iterations, the p-value for each trait was calculated by counting the instances in which the number of SNPs in the bootstrapped overlap was greater than or equal to the number of SNPs in the observed overlap.

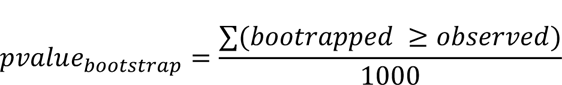

If *pvalue_bootstrap_* < 0.01, we assume that the observed relationship for that SNP is not random.

### Pathway analysis

Pathway enrichment analysis was completed using g:profiler and the Kyoto Encyclopedia of Genes and Genomes (KEGG) (Kanehisa et al. 2019), through the gprofiler2 R package. False discovery rate (FDR) multiple testing was used to select pathways with an adjusted p-value < 0.05 (supplementary table S6).

### Using deep learning to identify regulatory activity of SNPs associated with autism

To estimate the regulatory activity of SNPs associated with autism, the 2753 SNPs (autism-associated GWAS SNPs and those in LD) were queried into Sei, a deep learning algorithm for functionally annotating SNPs (Chen et al. 2022). Sei scored every SNP based on their predicted regulatory activity across 40 groups (Chen et al. 2022) (supplementary table S7). The SNPs were also separated into those that were spatially constrained adult and/or fetal eQTLs from those that were not (figure 1c). These groups were then compared based on their predicted regulatory activity and mean scores.

### Identification of traits that co-occur with autism within the New Zealand population

For robustness we compared the traits we determined to co-occur with autism with those seen in the New Zealand population. The Integrated Data Infrastructure (IDI) is a database maintained by Stats NZ that contains de-identified microdata about individuals in New Zealand (Statistics New Zealand 2022). The comoRbidity R package (Gutiérrez-Sacristán et al. 2018) was used to identify conditions that co-occur with autism within New Zealand’s hospital admissions data (31^st^ December 2015 −1^st^ January 2021). Conditions were based on the ICD-10 codes with F840 (childhood autism) and F845 (Asperger’s syndrome) used for autism. Within the comoRbidity package, Fishers exact tests were performed to calculate odds ratios for F840 and F845 occurring with all other ICD-10 codes. Odds ratios which were significant (corrected p-value after multiple testing ≤ 0.05) were selected.

### Ethics

Use of the Integrated Data Initiative within Stats NZ Data Lab was reviewed and approved by Statistics New Zealand (project number MAA2020-63) and the Auckland Health Research Ethics Committee (approval AH22495).

### Data access

Data analyses and visualisations were performed in R (version 4.2.0) through RStudio (version 2022.02.2). The Multimorbid3D pipeline, including bootstrapping, was performed in Python (version 3.8.8). Sei analysis was also performed on Python, using Jupyter Notebooks (version 6.3.0). Datasets and software used in the analysis are listed in supplementary table S9. All scripts are available on github (https://github.com/Catriona-Miller/Autism_Co-occurring_Traits).

## COMPETING INTERESTS STATEMENT

The authors have no competing interest to declare.

## Data Availability

All data produced in the present work are contained in the manuscript

https://www.ncbi.nlm.nih.gov/projects/gap/cgi-bin/study.cgi?study_id=phs001190.v1.p1

https://www.ncbi.nlm.nih.gov/sra?term=SRX2179249

https://www.ncbi.nlm.nih.gov/projects/gap/cgi-bin/study.cgi?study_id=phs000424.v8.p2

https://www.ncbi.nlm.nih.gov/projects/gap/cgi-bin/study.cgi?study_id=phs001900.v1.p1

https://www.ebi.ac.uk/gwas/

## ACKNOWLEDGEMENTS

We would like to thank the Genomics and Systems Biology Group (Liggins Institute, University of Auckland) for their helpful suggestions and discussions. Data from this work comes from the Genotype-Tissue Expression (GTEx) Project, which was supported by the Common Fund of the Office of the Director of the National Institutes of Health, and by NCI, NHGRI, NHLBI, NIDA, NIMH, and NINDS.

## AUTHOR CONTRIBUTIONS

CM performed data analyses, created visualisations, and wrote the manuscript. EG, JSW, and JJ co-supervised CM and commented on the manuscript. JOS co-supervised CM, conceived and directed the study, and commented on the manuscript.

## FUNDING

CM was funded by the University of Auckland Doctoral Scholarship. EG and JOS are funded by the Dines Family Foundation.

## Notes

### Competing Interest Statement

The authors have declared no competing interest.

### Author Declarations

The University of Auckland Human Participants Ethics Committee (UAHPEC) of the University of Auckland (Auckland, New Zealand) gave ethical approval for this work (Determining how SNPs contribute to Autism Spectrum Disorder (project number 024280)). Use of the Integrated Data Initiative within Statistics New Zealand Data Laboratory was reviewed and approved by Statistics New Zealand (project number MAA2020-63).

